# Assessment of psychological distress, coping, and spirituality in patients with resected, non-metastatic vs unresectable advanced cancer. Data from the NEOcoping and NEOetic_SEOM studies

**DOI:** 10.1101/2022.05.16.22275172

**Authors:** Veronica Velasco, Patricia Cruz-Castellanos, Raquel Hernandez, Adan Rodriguez-Gonzalez, Ana Fernandez Montes, Alejandro Gallego, Aranzazu Manzano-Fernandez, Elena Sorribes, Marta Zafra, Alberto Carmona-Bayonas, Caterina Calderon, Paula Jiménez-Fonseca

## Abstract

**Background:** Cancer negatively affects patients’ physical and mental health. This work sought to analyze the sociodemographic and clinical variables that modulate psychological distress in individuals with resected vs unresectable cancer before and after receiving systemic antineoplastic treatment, and to evaluate how different coping strategies and spiritual well-being influence psychological distress in these subjects.

**Methods:** Observational, prospective, consecutive study performed in 15 hospitals in Spain. The study consists of two cohorts: patients with resected locally and locally advanced cancer receiving adjuvant chemotherapy (NEOcoping study, 2016 and 2018) and those with unresectable locally advanced and advanced cancer, eligible for systemic treatment (NEOetic study, 2020 and 2021). Participants completed the following scales: BSI-18 (psychological distress) at baseline and after antineoplastic treatment; MINI-MAC (coping), and FACIT-sp (spirituality), before initiating systemic treatment.

**Results:** Fourteen hundred and fifty patients were recruited; 941 had resected cancer and 509 had advanced cancer. The advanced cancer sample included more males, older, less educated, and more unemployed individuals. Before starting systemic treatment, the advanced, unresectable disease group suffered more psychological distress than those with resected cancer (66.9 vs. 63.7, p=0.001) and used more coping strategies, especially positive attitude (79.1 vs. 75.6, p=0.001) and cognitive avoidance (64.3 vs. 53.6, p=0.001). Participants with resected cancer displayed greater spiritual well-being (36.5 vs 33.2, p=0.001). After receiving systemic treatment, psychological distress decreased in cases of advanced cancer and increased in resected cancer, albeit still higher in subjects with advanced cancer, particularly anxiety (61.4 vs 60.9) and depression (61.0 vs 60.6), (p=0.001 in both cases).

**Conclusion:** Patients with advanced cancer exhibit greater psychological distress, use more coping strategies, and have less spiritual well-being, but their psychological distress improves with antineoplastic treatment.

## Introduction

The incidence of cancer is on the rise, being 18.1 million in 2018 and estimated to reach 29.5 million in 2040 according to the World Health Organization (WHO). In Spain, the estimated rate of 280,100 cases in 2022 could rise to 341.000 in 2040 as per the Spanish Society of Medical Oncology (SEOM, for its acronym in Spanish). In addition to its high incidence, cancer has a negative impact on physical and mental health and a poor prognosis. Thus, despite all the diagnostic and therapeutic advances, it continues to be one of the leading causes of mortality, with a death toll of close to 10 million worldwide and 113,000 in Spain in 2020, second only to cardiovascular diseases. In 2040, cancer mortality is estimated to increase to 16.4 million worldwide and to 160,000 in Spain [1,2]

Given the aggressiveness and poor prognosis this disease entails, cancer patients are more susceptible to psychological disorders such as depression, present in up to 16% of all cases [3] or anxiety, existing in 38%, which adversely impact their quality of life [4]. Earlier meta-analyses, such as Wang Y-H et al from 2020 [5] conclude that psychological distress (symptoms of depression and anxiety) has been associated with higher cancer-specific mortality and worse survival.

Multiple factors play into to the development of these psychological symptoms, including how the individual reacts to their diagnosis, the fear of both disease-related symptoms and of treatment side effects, as well as living with the uncertainty of whether their tumor will progress or recur. Moreover, patients with social limitations have been found to manifest greater psychological distress; therefore, interventions focusing on social support can reduce distress in cancer patients, according to a 2014 meta-analysis [6]. Likewise, various psychological interventions can play a role in significantly alleviated cancer patients’ anguish, as concluded by the authors of a Chinese meta-analysis published in 2015 [7].

Coping is another important psychological aspect in this population, as it can be protective, by softening the negative impact of diagnosis, treatment sequelae, risk of relapse, and improve the socioeconomic and family sphere. Adaptive coping strategies (fighting spirit, positive reappraisal) can reduce some of the negative effects of cancer treatment, improve quality of life, and promote positive psychosocial adaptation to the events of the cancer process. In contrast, a person having a non-adaptive coping style, such as fatalistic coping, will be at greater risk for depression, isolation, and decreased quality of life [8,9]. Similarly, there is an inverse relationship between ones’ confidence in their ability to cope with cancer and distress, and a positive association between self-confidence and quality of life, as demonstrated by the meta-analysis by Chirico A et al [10]. Consequently, various psychological interventions must be conducted to promote better coping strategies that can modify cancer patients’ quality of life [11]. Individuals having a better feeling of psychosocial well-being have also been seen to cope better with the process of terminal illness [12].

Spirituality is the universal search for meaning and purpose in life, for connection with immaterial aspects of existence and transcendence. In their spirituality, people find a resource of strength that provides hope, comfort, and meaning to their life experiences, including cancer, in all its phases. Knowledge of prognosis, family and social support, autonomy, hope, and meaning to life contribute to positive psychospiritual well-being while emotional distress, anxiety, helplessness, hopelessness, and fear of death detract from psychospiritual well-being [13]. Being confronted for the first time with an incurable disease, at risk of recurrence/ progression, or the possibility of death itself leads some patients to pose a series of questions surrounding the meaning or purpose of existence. Thus, a Korean meta-analysis concluded that meaning-focused interventions might be a worthwhile way to improve end-stage quality of life for patients with advanced cancer [14]. Furthermore, spiritual interventions can lessen depression, anxiety, and hopelessness in people with cancer and provide physical and psychological benefits throughout all stages of the disease [12, 14]. When the correlation between religion/ spirituality and mental health in those with cancer has been examined, a positive relationship has been established [16], as well as an association with subjects’ ability to hold rewarding social roles and relationships [17]. Therein lies the importance for the medical team to consider the person’s spirituality when he/ she so desires, and for professionals with specific training in this area to intervene in the event of a spiritual crisis [18]. In that respect, a 2021 meta-analysis has confirmed that different psychosocial interventions are associated with small-to medium-sized effects on spiritual well-being among cancer survivors [19].

Given the importance of understanding the psychological impact of cancer and patients’ coping strategies, the aim of this study was to explore which biopsychosocial and clinical characteristics modulate the psychological state of individuals with resected or unresectable cancer before and after systemic antitumor treatment. We hypothesize that patients with advanced cancer (worse prognosis) are those with greater psychological distress. Additionally, we intended to study the effect of coping and spirituality on psychological distress in these patients.

## Materials and methods

### Patients and study design

This is an observational, prospective, consecutive study for which the data collected were obtained from two studies involving 15 hospitals in Spain. Individuals with a non-metastatic (resected) cancer came from the NEOcoping project, which is a study of the Continuing Care Group of the Spanish Society of Medical Oncology (SEOM) that was conducted between 2016 and 2018. Patients with advanced (unresectable) cancer were collected from the NEOetic project sponsored by the SEOM Bioethics Section between 2020 and 2021. Both studies were approved by the Ethics Committee of each institution and by the Spanish Agency of Medicines and Health Products, and all participants signed an informed consent form prior to inclusion.

The participants in each study were those who consulted in Medical Oncology during the interval between surgery for their neoplasm with curative intent (NEOcoping study) or between diagnosis (NEOetic study) and beginning of systemic treatment.

The population included comprised two groups of cases of histologically confirmed breast, colorectal, bronchopulmonary, or pancreatic cancer. The NEOcoping study enrolled patients with non-metastatic cancer that had been resected with curative intent and for which international clinical guidelines considered adjuvant systemic treatment to be an option. The NEOetic study included patients with advanced cancer (unresectable, stage IV and III) who were candidates for systemic therapy with non-curative intent. Individuals <18 years of age and those with any serious mental illness that would preclude them from understanding the study or who had any underlying personal, family, sociological, geographic, and/ or medical condition that might impede their participation were excluded. Another exclusion criterion was having previously received any systemic treatment for cancer or the only adjuvant treatment administered consisted of hormone therapy.

The study was based on a series of questionnaires that were explained to the patient by the medical oncologist; completed by the patient at home at the start of cancer treatment and handed in to the study assistants at the next visit. Each form contained detailed instructions and specified that its completion was voluntary and anonymous. The psychological distress questionnaire was also completed at the end of adjuvant treatment by patients with resected cancer (6 months after initiating treatment) and after the first radiological response evaluation study in subjects with advanced cancer (2-3 months after beginning antineoplastic treatment), a date closer to baseline, since this population has a worse prognosis and greater risk of early death.

### Measures and variables

The information was collected and updated by medical oncologists specially trained to meet the requirements of the study. Demographic and clinical data (age, sex, marital status, educational level, employment status, tumor location, and stage, and antineoplastic treatment received) were obtained directly from the patients and their medical records. Participants completed the following questionnaires: BSI-18 (psychological distress), MINI-MAC (coping), and FACIT-sp (spirituality). The version translated into Spanish and validated in previous studies was used [20] [19],[19], [20].

Brief Symptom Inventory (BSI). The BSI-18 consists of 18 items; the final score summarizes the respondent’s overall emotional adjustment or psychological distress over the course of the previous 7 days [23]. Each item is rated on a 5-point Likert scale, from 0 (not at all) to 4 (extremely). Cronbach’s alpha is between 0.81 and 0.90 [23]. This scale was validated in a Spanish-speaking population [21] and in the present study, it was evaluated at 2 time points, at the beginning and at the end of adjuvant treatment (series with a resected cancer) or after the first response assessment imaging study (series with advanced disease).

Mini-Mental Adjustment to Cancer (Mini-MAC). The Mini-MAC is a 29-item scale that gauges cancer-specific coping strategies as adaptive (cognitive avoidance, fighting spirit) or maladaptive (helplessness, anxious preoccupation, and fatalism) [22], [24]. When studying the psychometric properties of the scale translated into Spanish, we discovered a 4-factor structure which was used in this study and includes three subscales of the original questionnaire, helplessness, anxious preoccupation, and cognitive avoidance, plus a new subscale, positive attitude, which incorporates fighting spirit and the fatalism [22]. Each item is rated on a 4-point Likert scale ranging from 1 (definitely does not apply to me) to 4 (definitely applies to me). Cronbach’s alpha coefficients for each domain range from 0.62 to 0.88 [22], [24].

Functional Assessment of Chronic Illness-Spiritual Well-Being Scale (FACIT-sp). FACIT-Sp is a questionnaire consisting of 12 items rated on a five-point Likert-type scale ranging from not at all (0) to very much (4) and is organized into three subdomains, which address the components of spiritual well-being (meaning, peace, and faith) [25]. In the version translated into Spanish and other languages, the scale was divided into two factors, both of which were analyzed in this study, meaning/ peace and faith [20]. Internal consistency reliability coefficients ranged from 0.81 to 0.88 [25].

### Statistical analysis

Descriptive statistics were applied for demographic data and questionnaire responses. Absolute frequencies were used for categorical data and mean and standard deviation (SD) for quantitative data. Additional descriptive analyses were conducted by grouping patients by cancer type (bronchopulmonary, breast, colorectal, and pancreatic). Bivariate chi-square and t-tests were performed to examine differences between patients with resected, non-metastatic cancer and those with unresectable, advanced cancer with regard to sociodemographic, clinical, and psychological characteristics. A repeated measures analysis of variance was performed based on scores obtained at baseline and at 6 months (series with non-metastatic cancer) and 3 months (series with advanced cancer). Statistics were generated using a standard statistical software package IBM SPSS Statistics for Windows, version 23.0 (IBM Corp., Armonk, N.Y., USA).

## Results

### Sociodemographic and clinical characteristics

A total of 1535 patients were selected; 1450 were eligible for this analysis and 85 were excluded. Twenty patients failed to meet any inclusion criteria; 21 met one or more exclusion criteria, and 44 had incomplete data at the time of analysis.

Some 65% (n=941) of the cases had resected cancer distributed as 19% stage I, 36% stage II, and 45% stage III. Forty-five percent (n=509) had advanced cancer, 20% of which was stage III and 80% stage IV. Baseline sociodemographic and clinical characteristics can be found in Table 1. The mean age of patients with resected and unresectable cancer was 59.0 and 64.9 years, respectively. There were more men than women with advanced cancer; they were older and had a lower educational level than the cases of resected, non-metastatic cancer. The proportion of participants who were not working was higher in the group with advanced cancer, while among those with resected, non-metastatic cancer, most were employed.

**Table 1.** Demographic and clinical characteristics of patients (n =1450).

| Demographic and clinical characteristics | TOTAL<br>(n=1450) | Resected cancer<br>(n=941) | Advanced cancer<br>(n= 509) |
| --- | --- | --- | --- |
| Sex: n (%) |  |  |  |
| Men | 808 (56) | 235 (46) | 573 (61) |
| Women | 642 (44) | 274 (54) | 368 (39) |
| Age (years): mean (SD) | 61.0 (11.8) | 59.0 (12.2) | 64.9 (10.1) |
| Marital Status: |  |  |  |
| Married/ partnered: n (%) | 116 (78) | 714 (76) | 362 (83) |
| Educational level: n (%) |  |  |  |
| Elementary | 751 (52) | 507 (54) | 244 (48) |
| Intermediate | 699 (48) | 434 (46) | 265 (52) |
| Unemployed: n (%) | 887 (61) | 390 (41) | 497 (98) |
| Tumor: n (%) |  |  |  |
| Broncho Pulmonary | 190 (13) | 42 (5) | 148 (29) |
| Colon | 484 (33) | 398 (42) | 86 (17) |
| Breast | 355 (25) | 323 (34) | 32 (6) |
| Pancreas | 56 (4) | 16 (2) | 56 (11) |
| Type of adjuvant treatment |  |  |  |
| Chemotherapy | 912 (63) | 629 (67) | 283 (55) |
| Chemo- and radiotherapy | 327 (23) | 312 (33) | 15 (3) |
| Chemo- and immunotherapy | 49 (3) | 0 (0) | 49 (10) |
| Chemo- and targeted | 54 (4) | 0 (0) | 54 (11) |
| Others | 108 (7) | 0 (0) | 108 (21) |
| Death n (%) | 57 (6.9) | 21 (3.3) | 36 (17.8) |
| Abbreviations: *n* number, *SD* standard deviation |  |  |  |

The most common tumors were colorectal (33%) and breast (24%) in the complete sample, as well as in the group of patients with resected, non-metastatic cancer (42%, colorectal cancer and 34%, breast cancer, in this series). In participants with advanced cancer, the most common tumors were bronchopulmonary (29%) and colorectal (17%).

In patients with resected cancer with curative intent, postoperative treatment consisted of adjuvant chemotherapy, which was associated with radiotherapy in 33%. The most frequent treatment for subjects with advanced cancer was chemotherapy alone (55%) or combined with immunotherapy (10%) or targeted therapy (11%). Prior to completing adjuvant treatment (6 months) or the first response evaluation study (2-3 months), 6.9% of the sample had died, with a higher percentage of deaths in patients with advanced cancer (17.8%) than in those with resected cancer with curative intent (3.3%).

### Psychological adaptation to cancer

The differences in psychological adjustment by cancer type, resected, non-metastatic or unresectable advanced, are presented in Table 2.

**Table 2.**
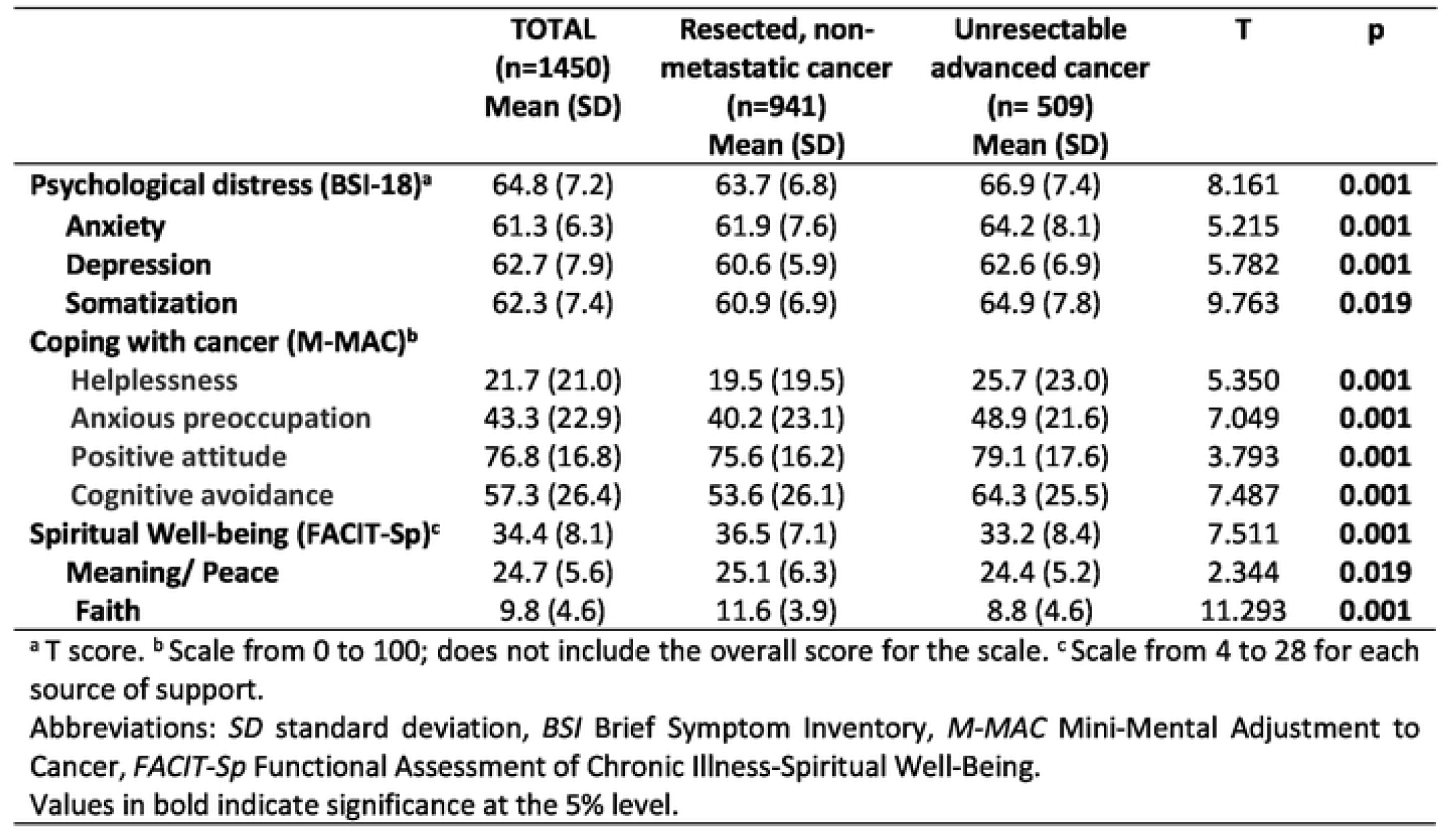
Differences in psychological distress, coping strategies, and spiritual well-being in patients with resected, non-metastatic vs unresectable advanced cancer.

|  | **TOTAL**<br>**(n=1450)**<br>**Mean (SD)** | **Resected, non-**<br>**metastatic cancer**<br>**(n=941)**<br>**Mean (SD)** | **Unresectable**<br>**advanced cancer**<br>**(n= 509)**<br>**Mean (SD)** | **T** | **p** |
| --- | --- | --- | --- | --- | --- |
| **Psychological distress (BSI-18)<sup>a</sup>** | 64.8 (7.2) | 63.7 (6.8) | 66.9 (7.4) | 8.161 | **0.001** |
| **Anxiety** | 61.3 (6.3) | 61.9 (7.6) | 64.2 (8.1) | 5.215 | **0.001** |
| **Depression** | 62.7 (7.9) | 60.6 (5.9) | 62.6 (6.9) | 5.782 | **0.001** |
| **Somatization** | 62.3 (7.4) | 60.9 (6.9) | 64.9 (7.8) | 9.763 | **0.019** |
| **Coping with cancer (M-MAC)<sup>b</sup>** |  |  |  |  |  |
| Helplessness | 21.7 (21.0) | 19.5 (19.5) | 25.7 (23.0) | 5.350 | **0.001** |
| Anxious preoccupation | 43.3 (22.9) | 40.2 (23.1) | 48.9 (21.6) | 7.049 | **0.001** |
| Positive attitude | 76.8 (16.8) | 75.6 (16.2) | 79.1 (17.6) | 3.793 | **0.001** |
| Cognitive avoidance | 57.3 (26.4) | 53.6 (26.1) | 64.3 (25.5) | 7.487 | **0.001** |
| **Spiritual Well-being (FACIT-Sp)<sup>c</sup>** | 34.4 (8.1) | 36.5 (7.1) | 33.2 (8.4) | 7.511 | **0.001** |
| **Meaning/ Peace** | 24.7 (5.6) | 25.1 (6.3) | 24.4 (5.2) | 2.344 | **0.019** |
| **Faith** | 9.8 (4.6) | 11.6 (3.9) | 8.8 (4.6) | 11.293 | **0.001** |

<sup>a</sup> T score. <sup>b</sup> Scale from 0 to 100; does not include the overall score for the scale. <sup>c</sup> Scale from 4 to 28 for each source of support.
Abbreviations: *SD* standard deviation, *BSI* Brief Symptom Inventory, *M-MAC* Mini-Mental Adjustment to Cancer, *FACIT-Sp* Functional Assessment of Chronic Illness-Spiritual Well-Being.
Values in bold indicate significance at the 5% level.

Psychological distress was greater among patients with advanced cancer with more symptoms of anxiety, depression, and somatization than those with resected cancer (p=0.001).

Of the four coping patterns identified, the one that scored highest in the total sample was positive attitude, while helplessness scored lowest. Individuals with unresectable advanced cancer scored higher on all four coping strategies than patients with resected, non-metastatic cancer (all, p=0.001).

Participants with resected cancer experienced more spiritual well-being in general, more meaning/ peace and faith, than those with advanced cancer (all, p≤0.019).

### Psychological change following antineoplastic treatment

We collected data pertaining to 627 patients with resected cancer and 166 who had completed adjuvant treatment (resected locally and locally advanced) or had completed the first response assessment study (unresectable advanced cancer) at the time of this analysis. Among the resected cancer group, 21 deaths were recorded; 86 patients abandoned chemotherapy prior to completing adjuvant treatment, probably because of associated toxicity (although this was not evaluated), and the final questionnaire could not be obtained for 207 patients. In the group of subjects with advanced cancer, 36 deaths were recorded and 473 losses to follow-up were recorded: 215 due to treatment withdrawal because of progression or clinical decline. Participants with resected, non-metastatic cancer had more somatic symptoms after completing treatment than those with advanced cancer (F_(1,791_)=29.480, p=0.001, η^2^=0.012) (Table 3). Depression lessened among individuals with advanced cancer following treatment, while in subjects with resected cancer it remained unchanged at baseline and at the end of treatment (F_(1,791)_=6.928, p=0.001, η^2^=0.009). Anxiety abated in both groups, albeit to a greater degree in patients with advanced cancer (F_(1,791)_=21.757, p=0.001, η^2^=0.027). Overall, psychological distress abated among participants with advanced cancer after 2-3 months of antineoplastic treatment, while it intensified in those with resected, non-metastatic cancer at the end of adjuvant treatment to coming close to equaling the score of patients with advanced unresectable disease at that time.

**Table 3.** Multivariate variance analysis of repeated measures before and after adjuvant treatment.

| Scales | Resected, non-metastatic cancer<br>(n=627) |  | t | p | Unresectable advanced cancer<br>(n=237) |  | t | p |
| --- | --- | --- | --- | --- | --- | --- | --- | --- |
|  | Pre Mean<br>(SD) | Post Mean<br>(SD) |  |  | Pre Mean<br>(SD) | Post Mean<br>(SD) |  |  |
| Psy. distress (BSI) | 63.8 (6.9) | 64.8 (7.1) | -3.651 | 0.001 | 66.2 (6.9) | 64.9 (6.8) | 3.308 | 0.001 |
| Anxiety | 61.8 (7.7) | 60.9 (7.5) | 3.375 | 0.001 | 63.5 (7.4) | 61.4 (7.3) | 4.633 | 0.001 |
| Depression | 60.5 (5.9) | 60.6 (6.2) | -0.465 | 0.642 | 62.3 (6.3) | 61.0 (5.9) | 3.781 | 0.001 |
| Somatization | 60.9 (6.7) | 64.6 (7.2) | -12.228 | 0.001 | 64.1 (7.2) | 64.5 (7.2) | -0.988 | 0.357 |

Abbreviations: *BSI* Brief Symptom Inventory, *Pre* before adjuvant treatment, *Post* after adjuvant treatment, *Psy* psychological, *SD* Standard Deviation.

## Discussion

This study is one of the few conducted in cancer patients that has focused on comparing the sociodemographic characteristics as well as the psychological distress, spiritual well-being and coping strategies of patients with different tumor stage and prognosis. On the one hand, these variables were analyzed in those with early cancer (resected with curative intent) and on the other hand in those with advanced cancer (unresectable or metastatic). In addition, we probed the impact of cancer treatments on the participants’ emotional state, for which psychological distress was analyzed prior to starting treatment and, again, after treatment.

In this study we found more psychological distress and greater use of coping strategies in patients with advanced unresectable disease and greater spiritual well-being in patients with resected disease with curative intent before starting antineoplastic treatment. After treatment, psychological distress decreased in patients with unresectable advanced cancer and increased in patients with resected, non-metastatic cancer. As for the clinical and demographic characteristics of the population studied, it is worth noting that the group with unresectable advanced cancer comprised more men, were older (around 65 years of age), had a greater proportion of unemployed, and had a primary school education. These patients had more lung, colon, and pancreatic cancer, and had received treatment predominantly with chemotherapy and chemotherapy combined with immunotherapy or targeted therapy and had a higher mortality rate. In contrast, the cohort with resected, non-metastatic cancer consisted of more women, younger individuals (around 59 years of age), diagnosed mainly with colon and breast cancer, who received chemotherapy alone or combined with radiotherapy in addition to surgery. These differences are based on the fact that breast cancer, which mostly affects women, is diagnosed at an average age of between 50-60 years (younger than other neoplasms) and thanks to screening programs, it is generally detected at earlier stages and, as such, is susceptible to both surgical and adjuvant treatment with curative intent. Lung and pancreatic cancers are usually diagnosed in advanced stages and lack early detection programs. The higher unemployment rate in patients with unresectable advanced cancer may be because they are older and, given the evolution of their disease, they may have a worse functional status. Moreover, in Spain, these patients are entitled to paid sick leave owing to their cancer.

This study confirms the hypothesis that, prior to commencing systemic treatment, individuals with unresectable advanced cancer experience greater psychological distress than those with early (resected) cancer. This may be the result of the greater impact the diagnosis might have on them because they suffer from an incurable disease with a worse prognosis and a lower survival expectancy [3] [26]. Nevertheless, following treatment, psychological distress, anxiety, and depression lessen among subjects with unresectable advanced cancer, probably because of better control of both the disease and its symptoms with antineoplastic therapy [27]. On the other hand, patients with resected, non-metastatic cancer suffered psychological worsening after completion of adjuvant treatment. This is possibly associated with treatment toxicity, of both chemotherapy and radiotherapy, as well as the sequelae of surgery, which tend to affect breast cancer patients, in whom changes in physical appearance interfere with social and work relations, triggering a decline in their psychological state [28] [29]. In addition, once adjuvant treatment has been completed, this group of patients is regarded as a cured with less follow-up, which may be accompanied by greater fear of disease recurrence that can impact on their psychological state.

When we analyzed the coping strategies used by our patients to adapt cognitively and behaviorally to the stressful situation posed by their cancer, more anxious worry, cognitive avoidance, positive attitude, and helplessness are observed among patients with unresectable advanced cancer, which is to be expected, given that they display greater psychological distress. In contrast, subjects with a resected cancer exhibited greater spirituality, meaning/ peace, and faith. Similar findings have been reported in previous studies in which greater spiritual well-being correlated with better cognitive and emotional functioning, in addition to less anxiety and depression [30]. It is worth noting that most studies to date that have probed into the influence of spiritual support in cancer patients have been carried out in the terminal or advanced stages of the disease [31,32]. As for the spirituality variables, meaning/ peace rated higher than faith, which may be attributed to the fact that spirituality transcends the traditional religious view, as evidenced in previous studies [33]. Thus, various meaning-focused pyschotherapeutic strategies have been proven to be instrumental in treating psychological, existential, or spiritual distress in patients with advanced cancer [34].

Previous research has corroborated the relationship between meaning/ peace and faith and coping strategies, anxiety, and depression, noting that optimism and social support foster enhanced spiritual well-being [35,36]. Similarly, the influence of spirituality/ religiosity on the different coping strategies has been appraised in patients with cancer. Spirituality has been revealed to be predictive of active coping [37]. Moreover, greater coping strategies have been associated with greater spiritual well-being, which entailed less negative, religious (maladaptive) coping and less psychological distress [38].

Other studies have examined specific series in Afro-American women with breast cancer in whom spirituality was seen to be the leading mechanism used to cope with their illness. In fact, efforts were made to promote spiritual interventions in nursing care [39]. Likewise, spirituality has been assessed in individuals with advanced disease in specific types of cancer, such as the study conducted among Norwegian patients with colon cancer who received palliative chemotherapy in which spiritual beliefs and the search for faith were seen to act as an inner buttress for the individuals [40]. Nevertheless, our study is the only one to compare the variables of spiritual well-being in subjects with different tumor stages and prognoses.

Few studies have analyzed and compared the biopsychosocial characteristics, coping strategies, and emotional status of individuals with early and advanced cancer. One study performed in the context of bladder cancer examined psychosocial stress in these two groups and found that patients with advanced disease displayed greater stress [41]. In this regard and in men with penile cancer, one study found that those who underwent surgery suffered greater psychological stress [42]. Another work in Chinese patients with different stages of breast cancer observed that low quality of life correlated with greater severity of somatic symptoms, depression, and anxiety [43]. A study was also conducted in patients with localized and advanced breast cancer to evaluate those factors that have a bearing on the development of chronic distress. The authors concluded that rumination, social constraints, and prolonged exposure to life stress increase the risk of developing persistent distress in advanced cancer [44]. However, most studies tend to focus on individuals with localized cancer, even if they present different tumor stages, as in the case of one study that demonstrated that good cognitive behavioral management decreased negative affect and intrusive thoughts in breast cancer patients who exhibited elevated postoperative distress [45]. Fatigue and emotional distress were also explored in a study of patients with stage I-III breast and colon cancer and highlighted that the participants who had completed treatment reported more symptoms, which is when they were given more advice and had more conversations with the physician [46].

Several studies have addressed patients with cancer at a similar stage of the disease, such as two Spanish projects, one of which analyzed coping strategies and psychological distress in patients with resected breast and colon cancer and found that they got worse after completing adjuvant therapy [28]. The other looked at coping and depression in subjects with different resectable cancers [3] and found that early psychological interventions, particularly in subjects with maladaptive coping strategies, can modulate the onset of depressive symptoms. Similarly, a study conducted in individuals with resectable digestive cancer that analyzed coping strategies and how they affect quality of life concluded that those patients with emotion-focused coping styles presented more deterioration on the quality of life scales [47]. Other studies have examined coping strategies without stratifying cases. Among them, one study involving Chinese patients with head and neck cancer looked at coping style after hospitalization and how it correlated with quality of life. The authors discovered that, after hospitalization, those subjects who had received treatments, such as radiotherapy, or those who had developed negative emotional coping styles (fatalistic, emotional, and avoidant) suffered a greater decline in their quality of life [48]. Another study conducted in an Iranian population that included mostly breast and colon cancer patients reported that quality of life correlated positively with avoidant coping style, while it correlated negatively with emotion-focused coping styles [49]. As for limitations, we ought to mention the differences in the sample size between the two series of cases studied. The sample with resected cancer was almost twice as large as the one with unresectable advanced cancer, probably because patients with more advanced disease present greater functional decline and are more difficult to recruit. In addition, the subjects with resected, non-metastatic cancer disease series were recruited over 3 years and those with advanced disease, over 2 years, coinciding with the COVID pandemic. Second, the follow-up sample is much smaller in unresectable advanced cancer than in resected, non-metastatic cancer. We cannot know how the loss of patients with advanced disease due to decline, progression, or early death has affected outcomes. Third, we must bear in mind that we have not performed a subgroup analysis that would enable us to ascertain how tumor location affects the variables studied. Fourth, although the study was controlled for clinical, sociodemographic, and psychological variables, the possibility cannot be ruled out that some other factor not contemplated may have played a role in the participants’ psychological distress. Finally, the questionnaires were completed by the patients themselves, which can lead to response bias resulting from interpretation errors, imprecise memory, or difficulty in understanding, and, despite proving useful to assess psychological distress, type of coping, and spiritual well-being, they should be used in conjunction with a clinical assessment.

As previously mentioned, one of the key contributions of our study and in contrast to most of those already performed to evaluate sociodemographic characteristics, coping strategies, spiritual well-being, and psychological distress, is that we have compared all these variables in participants with different tumor stages, grouping them into early cancer (resected cancer) and advanced disease (unresectable neoplasm) in a prospective study with a large sample size (n=1450).

In conclusion, our study illustrates that patients with unresectable advanced cancer experience greater psychological distress compared to those with resected, non-metastatic cancer. Consequently, the former use more coping strategies and have a lower level of spiritual well-being that allows them to cope with their suffering. They also comprise the group in whom psychological distress improves with antineoplastic treatment. Nevertheless, these results need to be confirmed in randomized clinical trials to promote psychosocial interventions focused on implementing coping strategies and spiritual interventions especially in individuals with advanced disease or resected, non-metastatic cancer who have completed adjuvant treatment to improve their mental health.

## Data Availability

All relevant data are within the manuscript and its Supporting Information files.

http://www.neocoping.es

https://www.neoetic.es

## Acknowledgements

The authors are grateful to the NEOetic and NEOcoping study researchers and the Bioethics Section and the Continuing Care Group of the SEOM for their contribution to this study. We would like to thank Priscilla Chase Duran for editing and translating the manuscript. The IRICOM team for the support of the website registry and specially Natalia G Cateriano and Miguel Vaquero.

## Authors’ contributions

V.V., C.C. and P.J.F developed the project, analyzed the data, and drafted the manuscript. The other authors recruited patients and provided clinical information, data curation, comments, and improvements to the manuscript. All authors participated in the conceptualization, interpretation and discussion of data, supervision, visualization, and the critical review and edition of the manuscript.

### Compliance with ethical standards

### Ethics approval

NEOcoping study was approved by the Research Ethics Committee of the Principality of Asturias (January 19, 2015) and by the Spanish Agency for Medicines and Health Products (AEMPS, for its acronym in Spanish) (April 14, 2015). NEOetic study was approved by the Research Ethics Committee of the Principality of Asturias (May 17, 2019) and by the AEMPS (May 8, 2019). The studies have been performed in accordance with the ethical standards of the 1964 Declaration of Helsinki and its later amendments. These studies are an observational, non-interventionist trials.

### Consent to participate

Signed informed consent was obtained from all patients.

### Consent for publication

Informed consent and approval by the national competent authorities includes permission for publication and diffusion of the data.

### Availability of data and material

Statistical analyses were performed with Statistical Package for Social Sciences (SPSS) software, 25.0 version (IBM SPSS Statistics for Windows, Armonk, NY: IBM Corp). The code is available upon request to the authors.

### Code availability

Patients are identified by an encrypted code known only to the local researcher. The code of the analyses is available upon request to the authors.

